# Identifying and preventing fraudulent responses in online public health surveys: Lessons learned during the COVID-19 pandemic

**DOI:** 10.1101/2022.12.12.22283381

**Authors:** June Wang, Gabriela Calderon, Erin R. Hager, Lorece V. Edwards, Andrea A. Berry, Yisi Liu, Janny Dinh, Amber C. Summers, Katherine A. Connor, Megan E. Collins, Laura Pritchett, Beth R. Marshall, Sara B. Johnson

## Abstract

Web-based survey data collection has become increasingly popular, and limitations on in-person data collection during the COVID-19 pandemic have fueled this growth. However, the anonymity of the online environment increases the risk of fraud, which pose major risks to data integrity. As part of a study of COVID-19 and the return to in-person school, we implemented a web-based survey of parents in Maryland, USA, between December 2021 and July 2022. Recruitment relied, in part, on social media advertisements. Despite implementing many existing best practices, the survey was challenged by sophisticated fraudsters. In this paper, we describe efforts to identify and prevent fraudulent online survey responses and provide specific, actionable recommendations for identifying and preventing online survey fraud. Some strategies can be deployed within the web-based data collection platform such as Internet Protocol address logging to identify duplicate responses and comparison of client-side and server-side time stamps to identify responses that may have been completed by respondents outside of the survey’s target geography. Additional approaches include the use of a 2-stage survey design, repeated within-survey and cross-survey validation questions, the addition of “speed bump” questions to thwart careless or computerized responders, and the use of optional open-ended survey responses to identify irrelevant responses. We describe best practices for ongoing survey data review and verification, including algorithms to simplify aspects of this review.

## Introduction

Web-based survey data collection has become increasingly popular, and limitations on in-person data collection during the COVID-19 pandemic have fueled this growth. Internet survey software and data capture systems (e.g., REDCap^12,13^, Qualtrics (Qualtrics, Provo, UT)) can reduce effort and expenditures associated with recruiting participants and may, in some applications, assist with accessing populations who may be difficult to reach via other means.^1,2,3^ Furthermore, the relative anonymity provided by online surveys may facilitate research involving marginalized communities or when respondents may otherwise be hesitant to disclose sensitive information.^4,5,6^

Despite potential benefits related to access, online survey research (particularly if offering incentives for completion) presents an increased risk of fraudulent activity as compared to face-to-face data collection. There is some evidence that fraud in research surveys has increased in recent years.^7,8^ While incentives can promote higher survey response and completion, they are also accompanied by an increased risk of interference from fraudulent responses.^2,9^ “Fraudsters” have various methods for finding surveys that involve incentives; for example, Meta (Facebook’s parent company) has an Ads Library that can help fraudsters find incentivized surveys that are advertised on their social media platforms, such as Facebook and Instagram. This resource can be exploited by fraudsters who may not be the intended target of a survey but may complete the survey solely for the incentive (“professional survey takers”) or utilize computer code to rapidly automate the completion of multiple surveys to receive multiple incentives.

Fraudsters pose a significant threat to research studies by undermining data quality and wasting resources.^6,10,11^ Past recommendations for mitigating online survey fraud have included tactics such as adding survey questions that reduce careless or automatic responding, implementing separate eligibility screeners, and implementing a Computer Automated Public Turing Test to tell Computers and Humans Apart (CAPTCHA) to outwit computer code that creates automatic responses.^10^ However, as fraudsters continue to fine-tune their tactics, existing recommendations may not adequately distinguish between legitimate and fraudulent respondents in public health surveys. Additionally, fraudulent respondents may attempt to outwit eligibility screeners by repeatedly testing combinations of responses to identify the combination that meets inclusion criteria and thereby grants them access to the survey (known in cybersecurity as a “brute force” approach). Once identified, fraudsters may complete multiple surveys using the same criteria. This can result in a disproportionate number of responses with identical or near-identical responses and poses a significant threat to validity should such tampering remain unidentified and unmitigated. Thus, as public health research and practice increasingly turn to web-based approaches, understanding and addressing threats to survey integrity are crucial.

In this paper, we describe specific strategies and lessons learned from efforts to identify and address fraudulent online survey responses as part of a mixed-methods study of parents’ perceptions of public health recommendations related to COVID-19 outbreak mitigation in schools in the USA.

## Materials and Methods

The Parents and Communities as Experts (PACE) Study sought to understand how parents of students in kindergarten through grade eight were navigating the return to in-person school following extended remote learning in eight counties in Maryland, USA. One element of the study was a survey implemented on the electronic data capture system, REDCap.^12,13^ Other study elements included a household mailed survey and focus groups. To be eligible, survey respondents had to be a parent or guardian of a public school student in grades K-8 during the 2021-2022 school year, live in one of eight counties in Maryland, USA, and read English or Spanish. The REDCap survey was fielded from December 2021 to June 2022. The Johns Hopkins School of Medicine Institutional Review Board approved the study protocol and respondents provided informed consent.

Participants were recruited through targeted social media ads on Facebook, community flyers and listserv postings, and postcard advertisements sent to households sampled to reach underserved and rural families. The Facebook recruitment strategy was developed based on strategies described by Ali et al. (2020), who recommended the use of a “single image” advertisement format designed to send social media users to the survey link. Recruitment materials referenced a $25 incentive for survey completion.^14^

Prospective participants followed a link to a public eligibility screener in REDCap. The eligibility screener was initially conceived simply to ensure respondents met eligibility criteria. The screener asked prospective respondents to indicate which of the eight target counties they lived in, confirm their status as a parent of a child in grades K-8 in a public school, and provide their zip code and how they heard about the survey so that recruitment could be monitored. Two initial survey security strategies were employed based on best practices for online survey security as outlined by previous work.^10^ First, a basic CAPTCHA was included in the screener to prevent access by fraudsters using automated software to complete survey responses *en masse* for incentives. Second, a REDCap feature that uses browser cookies to prevent duplicate responses was activated. Those eligible on the screener then received a personalized link to the survey via email. Survey responses were carefully reviewed for internal consistency and data quality by two study staff members. Below we detail the results of the initial fielding and iterative changes to the screener and survey to combat fraud.

## Results

The eligibility screener and survey were initially fielded on December 22, 2021, at 5:30 PM EST, and disseminated via targeted Facebook advertising. After the Facebook advertisements had been running for one hour—with a reach of only approximately 125 individuals—2,578 screeners were attempted and 950 responses to the survey were completed, highly suggestive of fraud. Among other suspicious indicators, responses came in batches of dozens per minute, overnight, and contained email addresses with strings of nonsensical letters and numbers. Whereas the advertisements were in the “Pending Review” stage (during which the advertisement was being reviewed by Facebook and not being delivered to members of the public) before 7:42 PM EST, 260 responses appeared to have been submitted before the campaign received approval and began delivery at 7:42 PM. This suggests that fraudsters may have located the advertisement prior to when public delivery began; it remains unclear how this would have been accomplished.

When the suspicious activity was detected in the REDCap survey at 11:33 PM on December 22^nd^, the Facebook advertisement campaign was deactivated. Strikingly, between the opening and closing of the campaign, only two “link clicks” on the advertisement were logged— while over 2,500 screeners were attempted. This suggests that the majority of response attempts were the result of the dissemination of the survey link through other channels.

We hypothesized that an individual or group of individuals may have posted the public survey link to the eligibility screener to online communities that share information regarding the manipulation of incentivized research surveys—a phenomenon that has been previously documented.^10^ It became clear that the screener was not adequately screening out fraudulent or suspicious respondents. As a result, the REDCap project was taken offline for redesign and Facebook advertisements were removed. Community-based recruitment and household mailings of recruitment postcards continued.

### Enhancements to eligibility screening

A revised REDCap eligibility screener that was harder to outwit was implemented in this new iteration of the survey fielding. For example, respondents were asked to choose their county of residence from a list of all counties in the state and to provide their zip code. Internal validation ensured that the participant lived in one of eight eligible counties and that their zip code was associated with an eligible county. Additionally, the respondent’s county of residence and zip code were required to be consistent between the screener questionnaire and the main survey if they were ultimately deemed to be eligible and received a survey link. A more complex CAPTCHA was added, as were two “speed bump” questions. Speed bump questions are designed to screen out bots and careless/automatic responders (an example of a speed bump question is, “The man couldn’t lift his son because he was so weak. Who was weak, the man or his son?”). Other types of speed bump questions include instructions for selecting an answer choice in the question stem that can be difficult to answer by automated scripts or rapidly responding fraudsters.^15^

### Time zone verification

On day 54 of survey fielding, to restrict access among respondents outside of the target area, a time zone difference calculation was implemented into the screener. This calculation compared the values of REDCap action tags “NOW” (i.e., the time reported by the client’s internet browser) and “NOW-SERVER” (i.e., the time reported by the institution’s REDCap server) to automatically calculate whether a respondent’s browser was set to a different time zone from the Eastern Time Zone—the time zone associated with the eligible study population. This calculation compared the client-side timestamp for screener completion with the server timestamp for receipt of responses. Respondents with greater than one hour difference were hypothesized to be attempting to complete the survey from alternate time zones and were thus barred from receiving the link for the study survey. Notably, however, we detected fraudulent respondents who avoided detection by intentionally setting their client-side time to match the time zone associated with the study (Eastern time), suggesting that at least some fraudsters may be aware of time zone disparities as a potential flag for suspicious activity.

The revised survey eligibility screener was deployed on January 26^th^, 2022. However, it became clear that our additional security enhancements did not sufficiently reduce the number of fraudulent attempts to complete the screener and many survey responses continued to be flagged as suspicious. Such flagged responses required additional review (for example, we triangulated responses to duplicate questions between the screener and survey to ensure internal consistency). Out of a total of 9,760 REDCap records, 978 had evidence of time zone disparities, and 316 respondents marked two or more counties in the screener when asked for their primary county of residence.

### Algorithms to screen for suspicious responses

To help screen potential responses for signs of fraud more efficiently, we implemented automated strategies deployed within REDCap to prevent the completion and submission of responses demonstrating clear signs of fraud. In addition, we developed a point-based methodology for manually identifying potential signs of fraud, derived from prior published guidelines.^6,8,11^ These manual methods involved detailed review and cleaning by study staff. Deriving guidance from the work of Ballard et al. (2019) we assigned points to observed indicators of suspicious activity, with point values corresponding to the degree that an indicator suggested fraud. Two or more points would cause a response to be marked as fraudulent. We have summarized the factors associated with varying point values in **Table 1**.

**Table 1:**
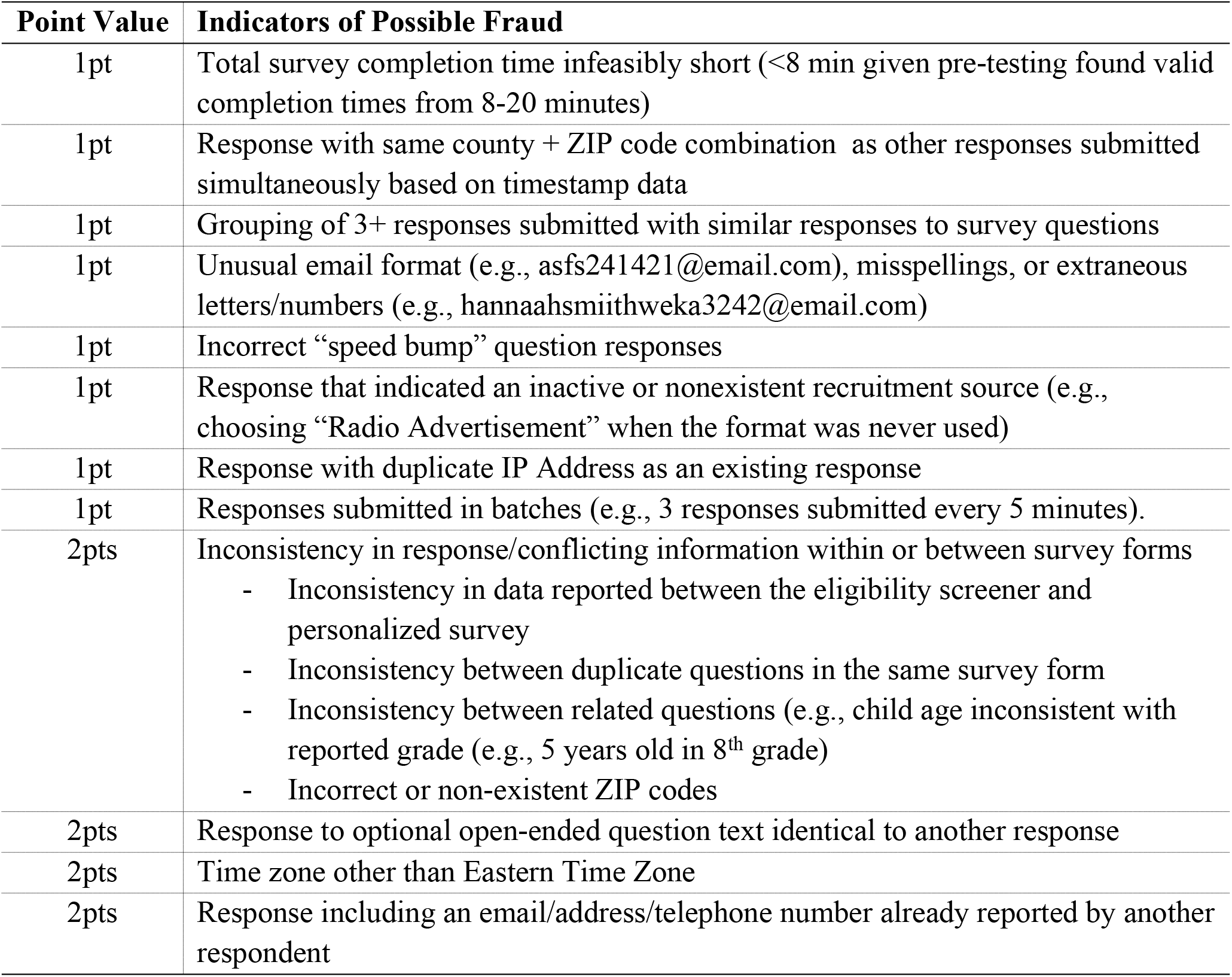
Example of points assigned to various indicators of fraud. (More points were an indication of greater risk).

### Internet protocol address logging

On day 75 of survey fielding, our institution’s instance of REDCap launched a form of anonymized Internet Protocol Address (IP) logging. Prior to this, due to privacy concerns, our instance of REDCap did not provide any IP logging or geolocation features. This newly added feature anonymized the respondent’s IP address (IP addresses are considered legally-protected personal health information (PHI) under the US Health Insurance Portability and Accountability Act (HIPAA)) but allowed us to identify multiple respondents from the same IP address. This additional feature allowed us to identify those who completed the survey more than once from the same IP. This feature was not used as rejection criteria but was rather utilized during the review of survey responses for data quality because two legitimate respondents from different households can share one IP address. It should be noted that this form of anonymized IP address logging does not permit the identification of fraudsters who use multiple IP addresses, such as via Virtual Private Servers (VPS), Virtual Private Networks (VPN), proxies, spoofed IP addresses, or methods of obfuscation.^8^ We considered duplicate IP addresses to be one point in our algorithm for quantifying the likelihood of fraudulent activity (Table 1).

### Creating duplicate survey databases

On day 126 of fielding, we created a copy of the REDCap web survey, with the same settings as the existing project, which was used specifically for Facebook recruitment directed toward Spanish-speaking respondents. This copied project was accessible to respondents via a different weblink. This copy presented numerous benefits for data integrity. We hypothesized the weblink to our original survey had been shared within covert networks of fraudsters.^10^ The use of surveys accessible through different weblinks may present an opportunity for researchers to limit the efficacy of such networks.

## Discussion

In fielding a survey of parents designed to capture perceptions about the safe return to school during the COVID-19 pandemic in the USA, we encountered a variety of threats to data integrity that required substantial time and effort to identify and address.

Mechanisms to prevent fraud should be an integral consideration during the survey design phase. Survey and database administrators should be aware of these mechanisms and recommend that all surveys implement measures to maintain data integrity and combat fraud. However, it is important to recognize that even existing recommendations can be circumvented by fraudsters with adequate resources. Automated measures to detect indicators of fraudulent activity and bar respondents demonstrating suspicious behavior from completing the survey can be “brute-forced” by fraudsters, who may attempt thousands of responses to determine a pattern of response that allows them to be deemed eligible. Upon determining such patterns, fraudsters will repeatedly utilize these strategies until they are detected. Here we recommend several strategies to limit inference and safeguard data integrity that are deployed 1) within the data collection platform, and 2) during the survey design phase.

### Strategies deployed within the data collection platform

Some strategies can be deployed directly through data capture or survey implementation platforms. First, consider omitting words like “survey,” “study”, or “research” from the text of survey platform weblinks to make them harder for fraudsters to find. Also, if possible, create multiple surveys associated with different web addresses. Cloned copies of the REDCap project allow researchers to rapidly change links as needed. Should the link for one survey instance be compromised by malicious actors, researchers can quickly pause the affected instance while permitting alternative instances to continue normal function. If available, researchers should activate IP logging, either direct IP logging or a HIPAA-compliant anonymized alternative that allows multiple responses from the same IP address to be flagged. (Note that even legitimate responses may contain duplicate IP addresses, as respondents may live or work where one IP address is associated with multiple households). If survey participants are restricted geographically, implement automatic comparison of client-side and server-side time stamps to identify responses completed outside of the target geography. However, it is important to know that fraudsters may adjust their computer clocks to circumvent these calculations.

### Strategies deployed in the survey design phase

Researchers should consider a two-stage process: an eligibility screener with a subsequent personalized survey link by email for those who are deemed eligible on the screener. Several design features can make it easier to reject or identify suspicious responses. In the screener, consider including a question with a false or impossible answer choice that could be used as a flag for suspicious activity. For example, a question about where participants heard about the survey could include “television ad” when no television ads were used. Add questions that are difficult for bots or computer algorithms to answer. For example, “speed bump” questions can help ensure respondents are reading the questions rather than answering indiscriminately.

An important way to identify suspicious activity is to validate the information respondents provide, within and/or across survey forms, depending on the study design. Validations within the same survey or screener can also be used to thwart “brute force” efforts to determine study eligibility criteria. For example, we asked participants to choose their county of residence from a list (including counties that were not eligible) and their associated Zip (postal) code. Counties and Zip codes were automatically compared and responses that did not match were deemed ineligible. Validations can also be used between two survey forms. For example, we asked participants to provide their county and Zip code on both the screener and again on the emailed personalized survey. If the information did not match, responses were flagged as suspicious. We also included the same question more than once on the personalized survey as a data quality/internal consistency check. While participants’ answers may vary for reasons unrelated to fraud, consistency is an important data quality indicator that can be tracked and monitored.

Finally, consider adding an open-ended response to survey forms such as “is there anything else you’d like to share with us on this topic?” Open-ended responses can be reviewed for nonsense or unrelated content. Multiple survey responses that include the same or similar text can also be used to identify responses completed automatically.

It is important to note that even if these suggested strategies are used, human monitoring and oversight are critical. Human oversight is needed to monitor responses for new or emerging threats to data integrity and to review activity flagged as suspicious to avoid inadvertently excluding eligible participants with valid responses. To avoid wasting research funds and further incentivizing fraudsters, our experience suggests that payment never be automated and should always include a human review. Other strategies such as confirming the identity of a participant with an email or phone call before releasing funds may also help but raise questions about ethical obligations to remunerate participants who are non-responsive to researchers’ efforts to contact them. These recommendations are summarized in Figure 1.

**Figure 1:**
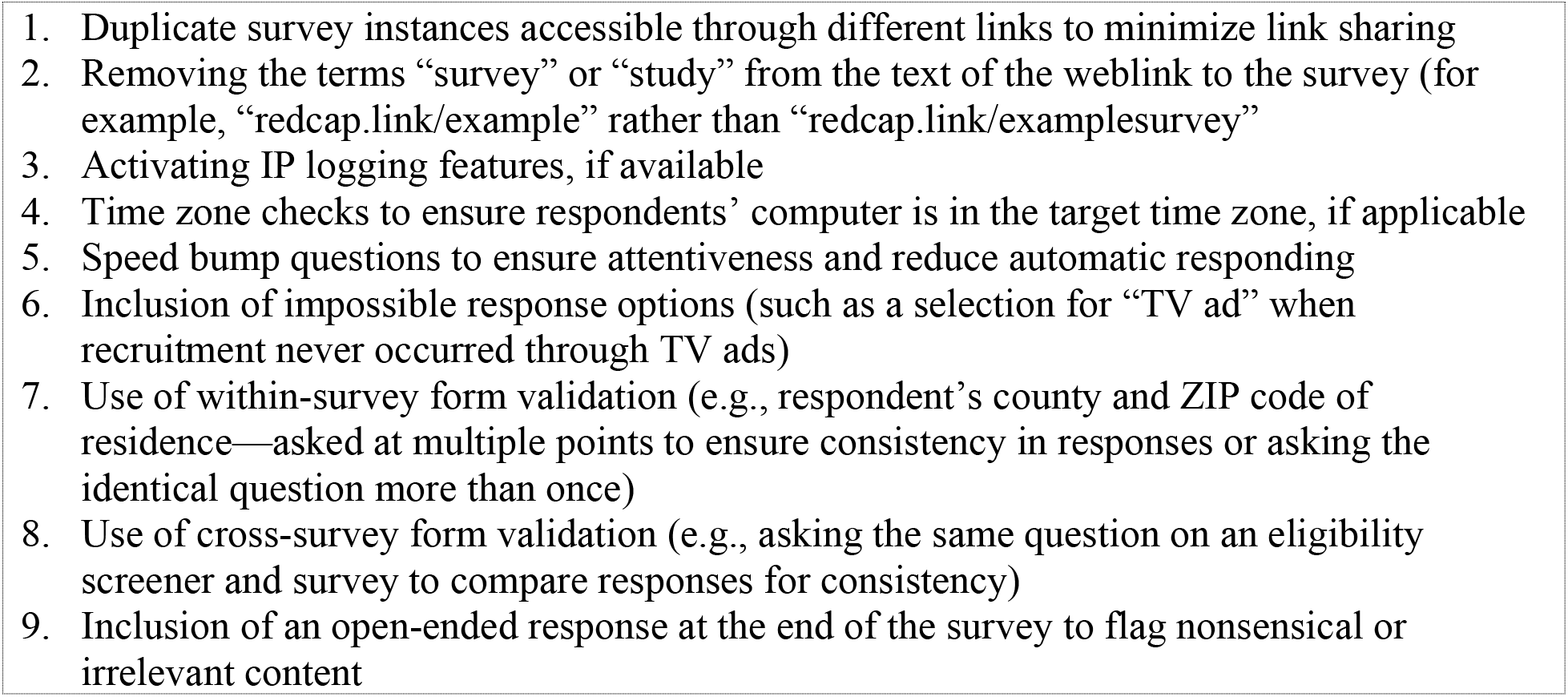
Summary of recommendations and best practices.

## Conclusions

Web-based research has become increasingly common in public health because it is comparatively inexpensive, less labor intensive than in-person data collection, and may expand access to a wider pool of potential research participants. Nonetheless, researchers must recognize threats to survey integrity posed by fraudsters who aim to gather study payments. Because fraudsters are constantly updating their approaches, continuous, ongoing monitoring is needed. Using strategies like screeners with embedded data quality questions and validation, time zone restrictions for completion, IP address logging, and multiple data forms can reduce threats to data integrity.

## Data Availability

The survey datasets generated and/or analyzed during the current study are available as part of the NIH RADx-UP Project. For more information regarding data availability, see https://myhome.radx-up.org/cdcc-resources/data-toolkit/

## References

1. Inan OT, Tenaerts P, Prindiville SA, et al. Digitizing clinical trials. npj Digital Medicine. 2020;3(1). doi:10.1038/s41746-020-0302-y

2. Pequegnat W, Rosser BRS, Bowen AM, et al. Conducting Internet-Based HIV/STD Prevention Survey Research: Considerations in Design and Evaluation. AIDS and Behavior. 2006;11(4):505–521. doi:10.1007/s10461-006-9172-9

3. Kayrouz R, Dear BF, Karin E, Titov N. Facebook as an effective recruitment strategy for mental health research of hard to reach populations. Internet Interventions. 2016;4:1–10. doi:10.1016/j.invent.2016.01.001

4. Das M, Ester P, Kaczmirek L. Social and Behavioral Research and the Internet: Advances in Applied Methods and Research Strategies. Routledge; 2018.

5. Thng C, Blackledge E, McIver R, Smith LW, McNulty A. Private sex workers’ engagement with sexual health services: an online survey. Sexual Health. 2018;15(1):93. doi:10.1071/sh16243

6. Ballard AM, Cardwell T, Young AM. Fraud Detection Protocol for Web-Based Research Among Men Who Have Sex With Men: Development and Descriptive Evaluation. JMIR Public Health and Surveillance. 2019;5(1):e12344. doi:10.2196/12344

7. Godinho A, Schell C, Cunningham JA. Out damn bot, out: Recruiting real people into substance use studies on the internet. Substance Abuse. 2019;41(1):3–5. doi:10.1080/08897077.2019.1691131

8. Pozzar R, Hammer MJ, Underhill-Blazey M, et al. Threats of Bots and Other Bad Actors to Data Quality Following Research Participant Recruitment Through Social Media: Cross-Sectional Questionnaire. Journal of Medical Internet Research. 2020;22(10):e23021. doi:10.2196/23021

9. Göritz AS. Incentives in Web studies: Methodological issues and a review. International journal of internet science. 2006;1(1):58–70.

10. Teitcher JEF, Bockting WO, Bauermeister JA, Hoefer CJ, Miner MH, Klitzman RL. Detecting, Preventing, and Responding to “Fraudsters” in Internet Research: Ethics and Tradeoffs. The Journal of Law, Medicine & Ethics. 2015;43(1):116–133. doi:10.1111/jlme.12200

11. Griffin M, Martino RJ, LoSchiavo C, et al. Ensuring survey research data integrity in the era of internet bots. Quality & Quantity. Published online October 5, 2021. doi:10.1007/s11135-021-01252-1

12. Harris PA, Taylor R, Thielke R, Payne J, Gonzalez N, Conde JG. Research electronic data capture (REDCap)—A metadata-driven methodology and workflow process for providing translational research informatics support. Journal of Biomedical Informatics. 2009;42(2):377–381. doi:10.1016/j.jbi.2008.08.010

13. Harris PA, Taylor R, Minor BL, et al. The REDCap consortium: Building an international community of software platform partners. Journal of Biomedical Informatics. 2019;95:103208. doi:10.1016/j.jbi.2019.103208

14. Ali SH, Foreman J, Capasso A, Jones AM, Tozan Y, DiClemente RJ. Social media as a recruitment platform for a nationwide online survey of COVID-19 knowledge, beliefs, and practices in the United States: methodology and feasibility analysis. BMC Medical Research Methodology. 2020;20(1). doi:10.1186/s12874-020-01011-0

15. Sonnad N. Easy questions that computers are terrible at answering. Quartz. Published August 2, 2016. Accessed November 3, 2022. https://qz.com/745104/easy-questions-that-computers-are-terrible-at-answering/

16. Singer E, Couper MP. Do Incentives Exert Undue Influence on Survey Participation? Experimental Evidence. Journal of Empirical Research on Human Research Ethics. 2008;3(3):49–56. doi:10.1525/jer.2008.3.3.

